# From Concept to Clinic: Real World Evidence for Autonomous AI Deployment in Primary Care Telemedicine

**DOI:** 10.64898/2026.03.18.26348749

**Authors:** Agustina Saenz, Elliot Schumacher, Dhruv Naik, Neal Khosla, Anitha Kannan

**Author notes:** Corresponding author: Anitha Kannan.

## Abstract

Systems powered by large language models are widely used for health information and advice, yet robust evidence for their safety and effectiveness in real-world clinical care remains lacking. Most existing studies evaluate general-purpose chatbots in artificial settings, failing to account for the critical role of system design, deployment context, and integrated safety mechanisms. Here, we report, to our knowledge, the first large-scale, clinician-blinded, real-world evaluation of a multiagent LLM-based system deployed within a nationwide U.S. primary care telemedicine platform, assessing readiness for task-specific autonomous deployment. In 2,379 real patient encounters, where users actively sought medical care and completed full visits with licensed clinicians, we compared the AI system’s intake diagnoses and disposition suggestions to those of treating clinicians, who were blinded to the AI’s outputs. The AI’s top-1 diagnosis matched the clinician’s diagnosis in 91.3% of cases overall, increasing to 96.3% among cases meeting a pre-specified safety confidence threshold, and 97.9% in common, lower-complexity conditions that met the same confidence threshold. Disposition accuracy was similarly high, with an overall error rate of 2.5% and no errors in suggestions to emergency room or home management. These results demonstrate that purposeful system architecture, rather than model capability alone, is essential for safe and effective autonomous clinical AI. We propose a staged, task-calibrated deployment framework, in which AI can be introduced autonomously for well-defined tasks with explicit safety gating and continuous monitoring, expanding scope as real-world evidence accrues. Our findings provide the first real-world evidence of readiness for safe autonomous clinical AI and offer a practical roadmap for its responsible deployment at scale.

## 1. Introduction

Approximately one in six U.S. adults now report using AI chatbots at least monthly for health information or advice^1^, and roughly one in four users of AI platforms submit a health-related prompt each week^2^. Users describe symptoms, seek guidance and act on recommendations, often without regulatory oversight and with very limited automated safety disclaimers.^3^ Large language models (LLMs) can now conduct multi-turn clinical dialogues^4^, elicit histories, and generate diagnosis and disposition recommendations^5–8^. They have also begun to demonstrate real-world utility in clinical workflows, such as facilitating primary-to-specialist care transitions^9^ positioning them not merely as documentation tools but as clinical decision-makers or assistive tools^10,11^ This is not a future scenario; it is current practice, and it is accelerating.

The evidence base for LLM performance in health care is growing, but much of it is derived from evaluations of systems, settings, or tasks that do not reflect clinical use in practice.

### Conflation of an LLM as a clinical system

many evaluations test a general-purpose chatbot interface^12–14^ and interpret the results as evidence about deployable clinical AI. Yet these evaluations are absent of key system elements such as clinical context integration, retrieval, safety guardrails, monitoring, and escalation pathways. This can lead to misleading conclusions about what is achievable when LLMs are embedded within purpose-built clinical systems.

### Simulation-to-reality gap

in controlled vignettes and simulations, LLMs can conduct diagnostic dialogues^4,15^, triage cases^16^, and support clinical decision-making^16^. More realistic simulated conversational evaluation frameworks have extended these assessments to multi-turn patient interactions.^15,17^. Approaches such as retrieval-augmented generation can further improve output quality^18,19^. However, these results may not translate to real-world use^20,21^, where performance is shaped by deployment constraints and user behavior (e.g., incomplete histories, ambiguity, time pressure, and workflow integration), not model capability alone.

### Task-in-isolation evaluation

most benchmarks focus on isolated tasks such as single-turn triage or diagnosis^22–24^ rather than end-to-end performance within a safety-critical workflow. Static evaluations, even when expert-curated, can miss requirements central to deployment, including iterative history-taking, uncertainty handling, modality-specific communication, and explicit safety escalation. For example, “chest pain” may warrant different questioning and next steps depending on whether it is reported via text, video, or in-person, and on the patient’s context; evaluating models in isolation fails to capture these dependencies that determine safe autonomous operation.

Taken together, these gaps leave unresolved whether LLM-based clinical systems can perform safely and effectively in real patient encounters rather than only in simulations, benchmarks, or narrowly standardized tasks. Previous FDA-authorized autonomous AI systems^25^, such as those for diabetic retinopathy screening,^26^ were developed for highly constrained use cases and validated under controlled conditions. Primary care presents a more demanding setting, characterized by heterogeneous presentations, incomplete histories, variable acuity, and less structured workflows. Rigorous evidence for autonomous AI in this context remains limited.

To our knowledge, this study reports the first large-scale, clinician blinded, real-world evaluation of multi-agent LLM-based system in a nation-wide US primary care telemedicine platform, assessing readiness for task specific autonomous deployment. In an analysis of 2,379 clinical encounters, the system demonstrated strong diagnostic concordance with treating clinicians, with the highest performance in lower-complexity cases and among encounters meeting prespecified safety thresholds, supporting autonomous deployment for selected well-defined tier-1 conditions when deployed with explicit safety thresholds and escalation pathways. Triage disposition performance similarly supported autonomous use for selected well-defined functions, including recommendations for emergency evaluation and home management, within a safety-gated workflow. Together, these findings support a staged, task-calibrated framework for autonomous AI deployment in health care, in which bounded functions with demonstrated real-world safety and reliability are introduced first and broader autonomy expands only as evidence accrues.

## 2. Methods

### 2.1. Study Design and Setting

To assess autonomous performance of our multi-agent system in routine primary care, we analyzed random samples of real-world encounters generated over an eight-month period on a nationwide U.S. telemedicine platform. The platform operates as a text-based clinical service for English-speaking adults across the United States, with clinician connection typically available within 15 minutes.

The study evaluated two deployed LLM-powered workflows on the same platform, accessed through different patient-facing entry points and analyzed separately. These workflows reflected different patient intents at entry: the symptom checker workflow was designed for users seeking AI-generated guidance about symptoms, self-care, and appropriate next steps, whereas the pre-visit intake workflow was used by patients who had already decided to consult a clinician and completed a multi-turn LLM-guided intake immediately before the visit. In both workflows, patients were informed that they were interacting with an AI-based system and could opt out or request clinician involvement at any time.

The **Symptom checker workflow** was used for suggesting dispositions (emergency care, home relief or virtual visit with clinician). In this workflow, the multi-agent system elicited symptoms and relevant history and presented a suggestion for how similar patients could think about next steps, namely emergency evaluation, home management, or continuation to a virtual visit.

The **Pre-visit intake workflow** was used for the diagnosis analysis. In this workflow, the multiagent system elicited symptoms, relevant history, and clinical context immediately before the clinician visit. Notably, the system’s diagnostic assessment was based solely on the intake conversation. During the subsequent clinician visit, patients may have provided additional information that was not available to the system.

### 2.2. Multi-agent Architecture

Our system consists of a multi-agent system in which LLMs serve as components within a broader orchestrated architecture designed specifically for primary care telemedicine. In contrast to general-purpose language models queried directly by patients, the safety and performance properties of this system reflect not only the underlying model capabilities but also the engineering and clinical design choices embedded throughout the architecture.

#### Multi-agent Orchestration and Prompt Development

The system coordinates multiple specialized agents, each responsible for a discrete clinical subtask within the patient encounter. Each agent is designed with a set of task-specific prompts developed by experts in both the clinical domain and LLM behavior. Prompts are scoped narrowly, capture the clinical reasoning required for the subtask, and explicitly address known LLM failure modes including prevalence bias and anchoring. An orchestration layer sequences these agents across the conversation, manages clinical context across turns, and enforces structured handoffs between subtasks. Prompt development followed a rigorous iterative process in which failure modes were characterized and addressed. Candidate prompts were evaluated against curated clinical cases, allowing for clear clinical validation. By treating prompt engineering as a form of clinical protocol development, we anchor all components in clinical best practices. The dataset analyzed in this paper was collected post-deployment and was not used in improving the system or prompts.

#### Safety Architecture

Safety is enforced through mechanisms operating in parallel throughout every step of the encounter rather than delegated to general model capability. When the features of an emergency issue are identified, a distinct system suggests that the patient uses emergency services or priority clinician connection. This separation ensures that safety-critical routing decisions are not dependent solely on model outputs and can be tailored to the clinical best practices of our clinic.

#### Clinical Reasoning and Disposition

Following history collection and safety screening, a clinical reasoning model generates a cumulative clinical assessment, including diagnostic output and dispositions. This assessment then undergoes a final independent validation check before informing the system’s subsequent recommendations and workflow decisions.

### 2.3. Dataset

#### Diagnosis dataset

The diagnosis dataset comprised 2,379 encounters among patients who completed the pre-visit intake and then proceeded directly to a virtual visit with a licensed clinician. This represented the predominant clinician-visit workflow on the platform and enabled evaluation at scale using an automated concordance framework. The dataset consists of encounters where the patients completed the multi-agent system intake in full, and is focused on a single clinical issue in which the treating clinician’s documentation reflected a sufficiently definite diagnostic impression for routine care, rather than multiple unrelated concerns or a broad, explicitly uncertain differential.

The system’s diagnostic assessment was based solely on its conversation with the patient. The clinician visit occurred immediately after the pre-visit intake, and patients may have provided additional information during that visit that was not available to the multi-agent system. Clinicians made independent diagnostic decisions informed by the AI–patient chat summary and their own direct interaction with the patient. Clinicians received no special instructions, were blinded to the system’s diagnostic conclusion, and practiced under their usual professional standards.

#### Disposition dataset

The disposition dataset comprised 161 encounters spanning three dispositions types: Emergency, home management and virtual care. Emergency and home management are substantially less frequent than virtual visit disposition on the platform, possibly because of the ability to access telemedicine after the symptom checker. Encounters in these categories of AI assigned emergency and home management were oversampled to enable meaningful safety evaluation. To ensure comprehensive coverage across all three categories, we constructed our dataset using a combination of two sources:

- AI-only encounters: Encounters in which patients did not proceed to a clinician after they got a suggestion of disposition from the symptom checker
- Clinician-reviewed encounters: Encounters where users proceeded to a virtual visit with a clinician. The clinician’s disposition became the reference standard

For AI-only encounters, we had three independent physicians first review the complete transcript of the AI-user interaction, including all questions asked and answers provided. Then, they evaluated whether all clinically relevant questions were asked, and if important context was missing, considered what additional information would have been necessary to make an accurate disposition decision. Based on the available information, and taking into account any missing but potentially critical context, each physician assigned the most appropriate disposition. All assessments were made independently, without knowledge of the other reviewers’ labels. The final reference-standard label assigned by majority voting. This approach ensures that the assigned label reflects the best possible clinical judgment, accounting for both the information present and any context that should have been sought during the encounter.

### 2.4. Diagnosis Performance Evaluation

For the diagnosis performance analysis, we compared the AI-generated diagnosis based on the intake conversation alone with the treating clinician’s final diagnosis using a structured concordance rubric adapted from the clinician evaluation framework described by Tu et al.^4^ (Appendix). The adapted rubric assessed degrees of clinical alignment on a five-point scale ranging from unrelated to exact match, rather than using a binary correct-or-incorrect judgment. We modified the instructions to better capture clinically correct diagnoses that differed in wording, diagnostic breadth, or implied specificity but referred to the same or a clinically equivalent condition, thereby reducing false discordance due to terminology alone. We used GPT-4.1 to apply the adapted rubric and assign concordance ratings to AI–clinician diagnosis pairs at scale.

Exact match was defined as the top-1 AI-generated diagnosis being identical or clinically equivalent to the treating clinician’s diagnosis, accounting for commonly implied characteristics and equivalent terminology, for example, treating “common cold” as equivalent to “upper respiratory infection” or inferring bacterial etiology where clinical standard. Extremely relevant was defined as a top-1 diagnosis that was very close but not an exact terminological match. Concordance was prespecified as a rating of either exact match or extremely relevant. Both categories were considered concordant because the reference standard was the treating clinician’s diagnosis as documented in routine care rather than under standardized study-specific labeling rules. Clinicians were not instructed to document diagnoses in a uniform way for research purposes, and therefore the same clinical condition can be recorded with different words or at different levels of granularity. A strict exactmatch criterion would therefore introduce false discordance by treating routine variation in clinical documentation as diagnostic disagreement. Representative examples of diagnosis pairs rated as extremely relevant are shown in Appendix Table 1.

**Table 1:** Examples of Diagnoses Considered Extremely Related.

| **AI Diagnosis** | **Clinician Diagnosis** |
| --- | --- |
| Traumatic nail injury (nail avulsion of finger) | Injury of nail bed of finger |
| Acute mechanical back pain | Acute mid to upper back pain |
| Mucocutaneous candidiasis | Candidal vulvovaginitis |
| Acute viral rhinosinusitis | Acute sinusitis |
| Acute viral upper respiratory tract infection | Acute viral rhinosinusitis |
| Acute bacterial skin and soft tissue infection of finger | Laceration with possible bacterial infection |

Treating clinicians had access to the AI-generated subjective documentation (a structured summary of the patient’s reported symptoms and history), but did not see the AI-generated diagnosis and treatment plan prior to the clinical encounter. Patients then completed a clinician visit, typically within 15 minutes of AI intake, during which the clinician could obtain additional history through direct chat. The multi-agent system did not have access to that subsequent clinician-patient conversation. This design reduced the risk of anchoring bias in the reference standard and strengthened the validity of the concordance analysis. Diagnostic accuracy was defined as concordance between the AI-generated diagnosis at the end of the intake dialogue and the primary diagnosis documented by the treating clinician at the end of the encounter. In cases of diagnostic discordance, errors were further characterized to distinguish between reasoning failures, incorrect conclusions drawn from available clinical information, and information-gathering failures, defined as insufficient elicitation of clinically relevant history during the intake dialogue.

### 2.5. Disposition Accuracy Evaluation

Disposition accuracy was evaluated by comparing the system-suggested disposition with the reference-standard label assigned as described in Section 2.2 across 3 categories: emergency evaluation, home management, and virtual visit. A suggestion was considered correct only when it exactly matched the reference-standard label. We report overall accuracy as well as concordance for each disposition category. Because the clinical significance of discordance depends on its direction, errors were further classified as undertriage when the AI suggested a lower-acuity disposition than the reference standard and as overtriage when it suggested a higher-acuity disposition.

## 3. Results

### 3.1. Diagnosis Performance

Overall, top-1 diagnosis concordance was 91.3% across all 2,379 encounters. A substantial number of encounters were flagged for clinician review by our multi-agent system as a precaution in the setting of diagnostic uncertainty rather than known error. Of the 2,379 encounters, 1,094 met the prespecified diagnostic-confidence threshold. In this subset, top-1 diagnostic concordance was 96.3%, as compared with 87.0% among the remaining 1,285 encounters. The threshold preferentially excluded diagnostically weaker outputs, including 82.1% of diagnoses adjudicated as unrelated (55 of 67) and 86.1% of those adjudicated as somewhat related (62 of 72).

We next evaluated performance in a prespecified subgroup of lower-complexity, high-frequency tier-1 conditions, including uncomplicated urinary tract infection, vulvovaginal candidiasis, bacterial vaginosis, upper respiratory infections among others. We applied the same diagnostic-confidence threshold as before. In this subgroup, top-1 diagnostic concordance was 97.9% (n=523) (Fig. 2). Concordance was 96.4% for URI/acute respiratory symptoms (n=303), 93.5% for bacterial vaginosis (n=31). Concordance was 100% for uncomplicated cystitis (n=172), yeast vaginitis (n=53) and each of the remaining tier-1 strata. (n≤11 each).

**Figure 1:**
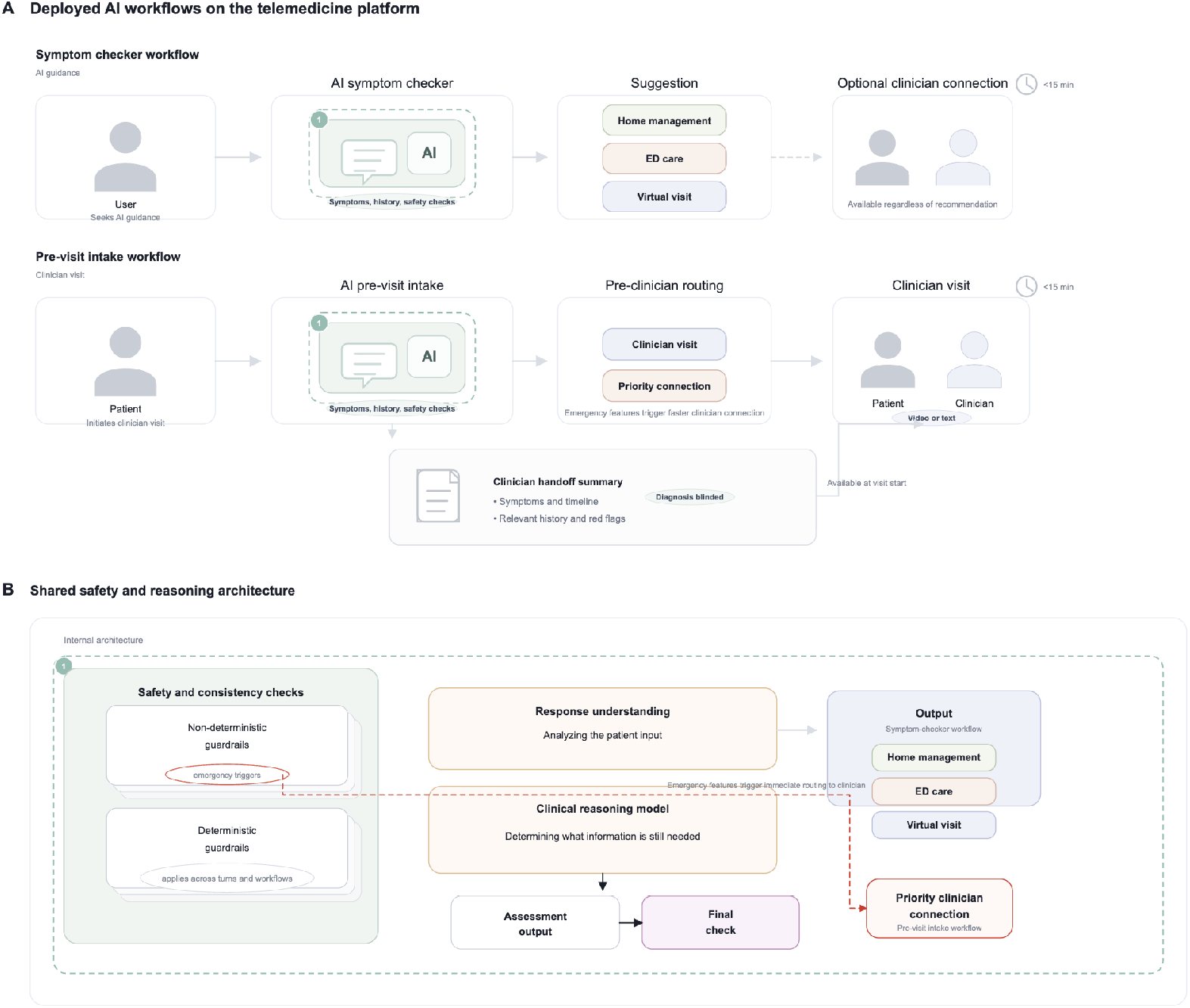
Deployed AI workflows on the telemedicine platform and shared system architecture. Panel A shows the two patient-facing AI workflows evaluated on the platform. In the symptom checker workflow, users seek AI guidance and receive a suggestion for home management, ED care, or a virtual visit, with an optional clinician connection available regardless of the suggestion. In the pre-visit intake workflow, patients initiating a clinician visit complete an AI intake immediately before the visit; emergency features can trigger priority clinician connection. In both workflows, a subjective summary is generated and made available at visit start, with the AI diagnosis and disposition blinded. Panel B shows the shared safety and reasoning architecture underlying both workflows. Each patient message passes through deterministic and non-deterministic safety and consistency checks, followed by response understanding and clinical reasoning. The system then produces an assessment output that undergoes a final check, while emergency triggers are monitored continuously and can prompt immediate priority clinician connection.

**Figure 2:**
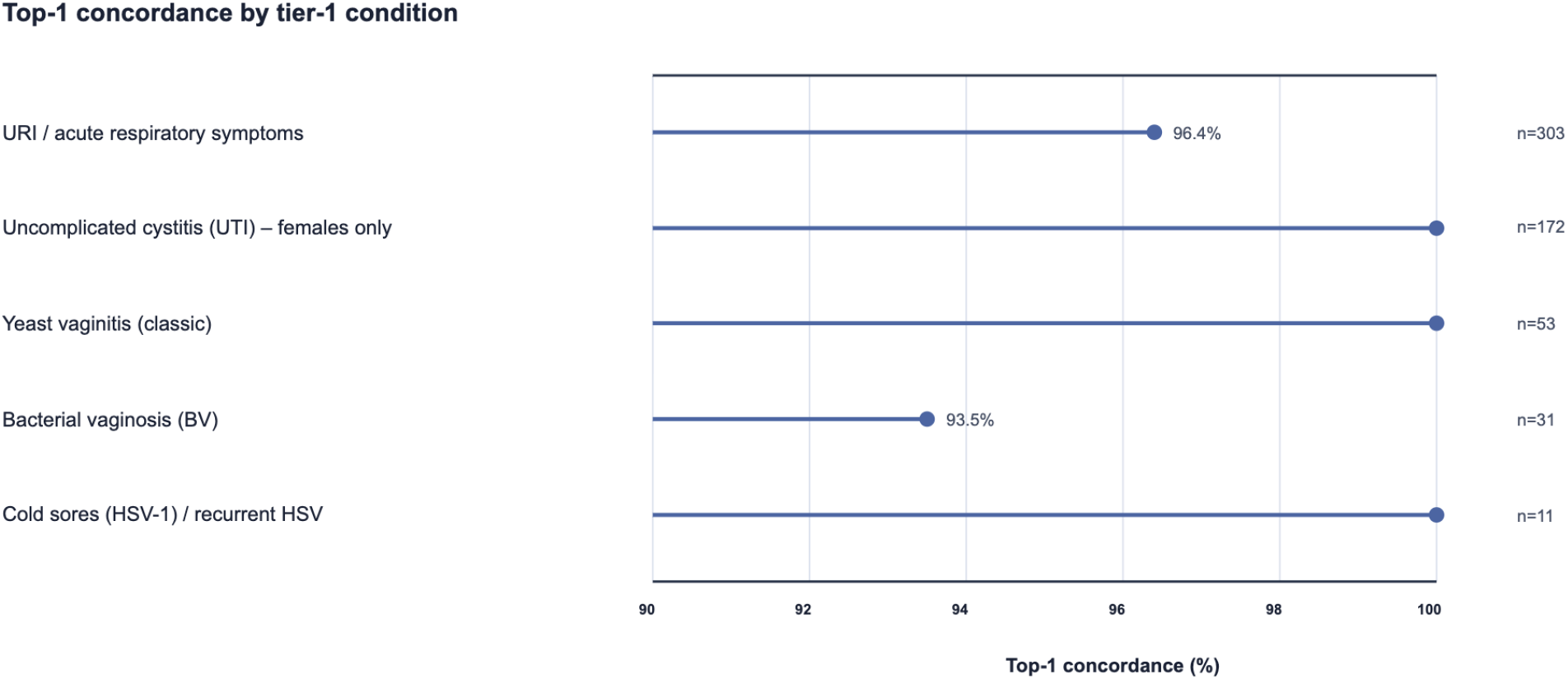
Top-1 diagnostic concordance by condition among encounters passing the system’s prespecified confidence criteria. Concordance was high across all tier-1 conditions, reflecting consistent performance in well-defined, lower-complexity primary care presentations.

### 3.2. Disposition Accuracy

Of the 161 encounters in the disposition dataset, we found an overall error rate of only 2.5%. Among the 25 cases in which the multi-agent system suggested home management and the 76 in which it suggested emergency evaluation, concordance with the reference standard was 100%. Because patients receiving these suggestions may not proceed to clinician review within the platform, errors in these categories may not be intercepted by our clinicians, making high concordance particularly important. All four discordant cases occurred among the 60 cases in which the system suggested a virtual visit. Precision for virtual visit suggestions was 93.3%, indicating that discordant cases represented a small minority of this group. Incorrect virtual visit suggestions could still be corrected by clinicians within the platform, providing an additional safety check and an opportunity for redirection.

Of the 4 discordant virtual-visit cases, 3 were adjudicated as requiring emergency evaluation despite the AI’s earlier virtual-visit suggestion. In the remaining case, the system suggested a virtual visit, whereas the treating clinician determined that the symptoms could be managed at home without prescription or intervention.

## 4. Discussion

This study provides real-world evidence of a multi-agent system operating across the full disposition and diagnosis workflow of a primary care telemedicine platform serving patients across the United States. Two findings stand out. First, diagnosis accuracy was 96.3% for diagnoses approved to be surfaced to patients, rising to 97.9% for lower complexity, high-frequency conditions reflecting a system that defers to clinicians when diagnostic confidence is insufficient rather than surfacing uncertain conclusions. Second, disposition accuracy was high and compared favorably against both human and digital benchmarks. Notably, this is the first large scale, clinician-blinded, study to measure diagnostic capability of multi-agent system in a live clinical environment, demonstrating that the system is rapidly approaching, and in some cases saturating, the performance levels typically expected of physicians. Together, these findings support autonomous operation for selected well defined primary care tasks within a safety-gated workflow, without requiring real-time clinician review for every encounter.

### 4.1. Diagnosis Performance in Context

An overall diagnosis concordance of 91.3%, or 96.3% with the safety mechanism, is much higher than any published results, both those based on simulation or vignettes studies or through prospective study. In a previous study^20^, models succeeded 94.9% of the time under ideal, controlled conditions. In contrast, when tested with real patients, the same models identified relevant conditions in fewer than 34.5% of cases. In another recent prospective study, the top-1 diagnosis accuracy was 61%^21^ while their initial simulation based studies reported much higher top-1 accuracy.^10^

In the clinician-led virtual-visit workflow, the system’s top-1 diagnosis concordance with the treating clinician was high overall and increased further among encounters that passed the pre-specified confidence gate. The gain is attributed to the architecture, not just to the underlying model, reinforcing the principle that clinical AI should be evaluated as a system rather than in isolation. Notably, prompt-based automated concordance assessment does not fully capture documentation limitations where clinicians occasionally recorded presenting symptoms rather than formal diagnoses, or diagnosis without concurrent conditions, suggesting that true concordance is possibly higher than reported.

For lower-complexity, higher-frequency tier-1 conditions like uncomplicated UTI, vulvovaginal candidiasis, and cold sores among others, concordance reached 97.9% supporting potential autonomous deployment. These are well defined diagnoses with standardized management guidelines. As real world evidence accumulates, the scope and level of diagnostic autonomous diagnosis is expected to expand.

Among tier-1 encounters that met the confidence threshold, we manually reviewed the 11 cases that were flagged as likely correct by the system but did not achieve concordance with the treating clinician diagnosis. Four reflected disagreement, including one encounter in which the patient’s history changed between intake and the clinician visit (reporting a fishy odor and gray discharge to the AI but later denying discharge and instead reporting cloudy urine) and one in which the system labeled allergic rhinitis while the clinician labeled a viral upper respiratory infection, with similar symptomatic treatment. Two reflected differences in diagnostic specificity (for example, “upper respiratory infection” versus a specific viral etiology such as COVID-19 or influenza). Two reflected safety-oriented severity framing (for example, urinary tract infection versus “complicated UTI” when flank pain with nausea, chills, or similar features were reported). The remaining were clinically aligned but differed in wording or documentation emphasis rather than representing distinct conditions. Overall, these reviews suggested that most residual discordances in tier-1 conditions reflected labeling or safety-framing differences rather than clinically meaningful disagreement.

### 4.2. Disposition Performance in Context

An overall disposition error rate of 2.5% compares favorably against published benchmarks. In primary care settings, under-triage rates of 10–19% and over-triage rates of 13–19% have been reported, with even wider variation in emergency settings.^13^ Digital symptom checkers, the closest analogous technology, have demonstrated mean error rates of approximately 50%, with individual tools ranging from 22–72%.^27^ In another study, 51.6% of true emergencies were under-triaged, particularly when emergency status depended on clinical trajectory rather than classic presentation.^14^ Direct comparison is limited by differences in triage / disposition definitions, patient populations, and error classification across studies, and should be interpreted accordingly. Nonetheless, the directional finding is consistent across every metric: the system’s error rate fell below the lower bound of human primary care disposition error rates as well as substantially below digital symptom checker performance.

Three undertriage errors were identified (1.9%), all involving patients suggested for virtual visits who required urgent or emergency evaluation. Critically, when multi-agent system suggested patient for emergency evaluation, it was not overtriaging them and similarly, when it suggested for home management, not undertriaging them. The absence of errors across 76 emergency dispositions and 25 home management dispositions is a particularly important safety signal given the consequences of errors in these categories.

The three undertriage cases were also surfaced through the system’s real-time safety monitoring process, illustrating a broader principle: that autonomous deployment accompanied by continuous output validation identifies patterns that controlled evaluations and retrospective benchmarking cannot surface. Unlike reactive safety models that detect failures after they reach patients, this system anticipates potential red flags, proactively guides outputs, and catches errors before they occur. This represents a model for responsible autonomous AI deployment that the field has described theoretically but rarely demonstrated empirically: a system that continuously checks its own work, recommends modifications when it senses risk, and defers to a clinician whenever uncertainty is detected.

## 5. Toward Calibrated Autonomy: A Framework for Deployment

The case for autonomous AI in primary care extends beyond performance. Average wait times for a primary care appointment now exceed 31 days across major US cities, with some markets averaging over two months^28^. For patients who cannot access timely care, autonomous AI is not a convenience but a necessity. A system that directs the patient safely, recommends home management when appropriate, and connects patients to a clinician when needed, at any hour, without an appointment, addresses a structural gap the healthcare system has failed to close. The question is no longer whether patients will use autonomous AI for medical guidance; it is whether that AI will be built with the safety standards they require.

The path from AI-assisted to fully autonomous clinical care needs to mirror a familiar model in medical training. A new resident requires supervision for complex diagnostic decisions but can safely recommend symptomatic management for a self-limited illness without attending oversight from day one. As competence is demonstrated across a broader range of tasks, supervision requirements decrease, not because oversight is abandoned but because it is calibrated to the evidence. Autonomous clinical AI should follow the same logic: beginning with well-defined, lower-acuity tasks where the evidence base is strong, operating asynchronously with available clinician review, and expanding to fuller autonomy as real-world data accumulates.

The findings presented here support this directly. Autonomous AI readiness is not a binary threshold but a task-specific determination. The appropriate framework is not autonomy versus oversight but calibrated autonomy: matching the level of autonomous function to the evidence base for each specific task or diagnosis, with mandatory prospective monitoring and defined governance. For bounded tasks such as directing patients to the appropriate level of care, including emergency evaluation or home management, and for selected tiered diagnoses, the error rates observed here support autonomous AI-generated guidance without mandatory real-time clinician involvement in every encounter. This has direct implications for clinician capacity: such a calibrated autonomy with effective monitoring system that performs at or above human accuracy frees clinician time for encounters where judgment is irreplaceable, while extending access to patients who might otherwise forgo care entirely. Because care was delivered entirely via chat across a geographically diverse US patient population, these findings are better reflective of performance across a broad range of health literacy levels, socioeconomic backgrounds, and geographic settings including rural and underserved communities frequently underrepresented in single-institution or simulation-based clinical AI evaluations.

Current FDA authorization pathways were designed for narrow, static diagnostic tools and are ill-suited to generative AI systems that operate across broad clinical tasks and evolve continuously. Rather than applying legacy frameworks not designed for this technology, regulators should develop pathways that recognize real-world performance data, prospective safety monitoring, and iterative improvement as valid evidence of safety and efficacy.

Evaluating a model and evaluating a clinical system are not the same thing, a distinction the field must make explicit as autonomous AI research expands. Studies assessing frontier or open-source models deployed without safety architecture, monitoring layers, or clinical guardrails measure model performance in isolation^29^. A model that fails in that context may perform safely when embedded within a purpose-built system designed to anticipate errors, validate outputs, and defer to a clinician when needed. Findings from model-only evaluations and system-level evaluations should not be reported interchangeably if the field is to build a reliable evidence base for deployment decisions.

Autonomous AI encounters also expose a gap in existing liability frameworks. Traditional malpractice doctrine assigns liability to a licensed practitioner; when no practitioner is involved, that framework has no clear application. Rather than treating this as a barrier to deployment, it should be treated as an opportunity to develop purpose-built liability frameworks and patient disclosure standards that reflect how this technology actually works and that enable rather than obstruct its responsible expansion.

Based on the evidence presented here and the broader trajectory of the field, we propose an initial framework for responsible deployment in primary care (Fig 4).

**Figure 3:**
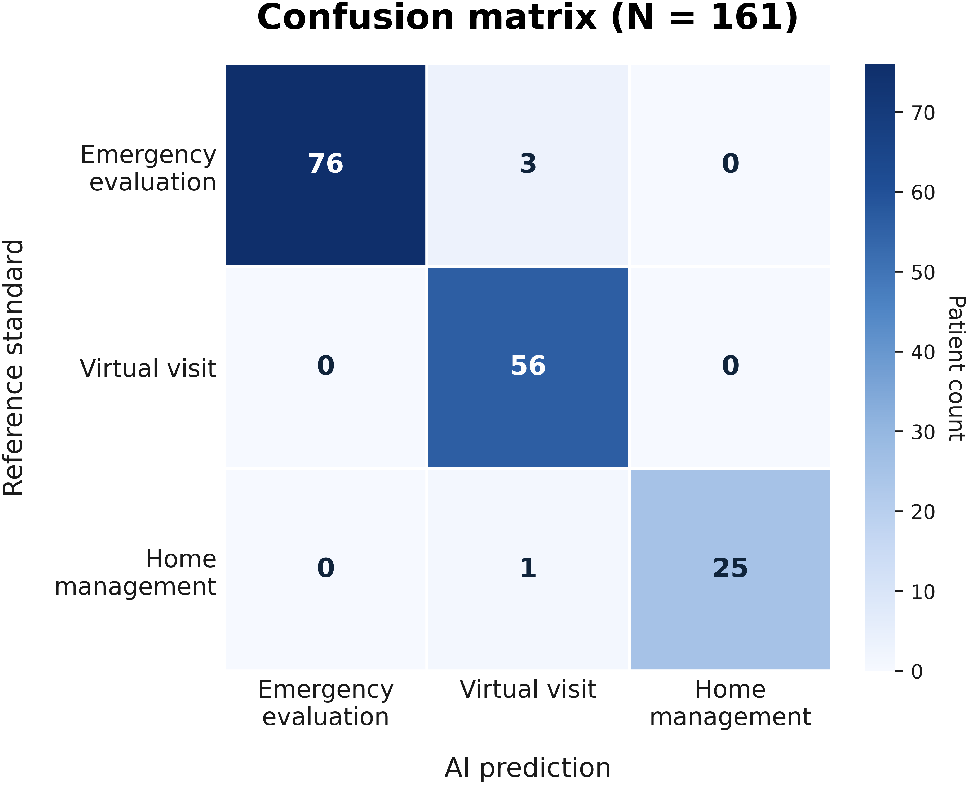
We compare the AI-generated dispositions against the physician reference standard. The overall disposition error rate was 2.5% (4 of 161). For emergency and home care, the AI system achieved 100% concordance. This is critical for patient safety as they will not be directed to see a clinician on our platform. Among cases for which the AI suggested a virtual visit, concordance with the reference standard was 93.3%.

**Figure 4:**
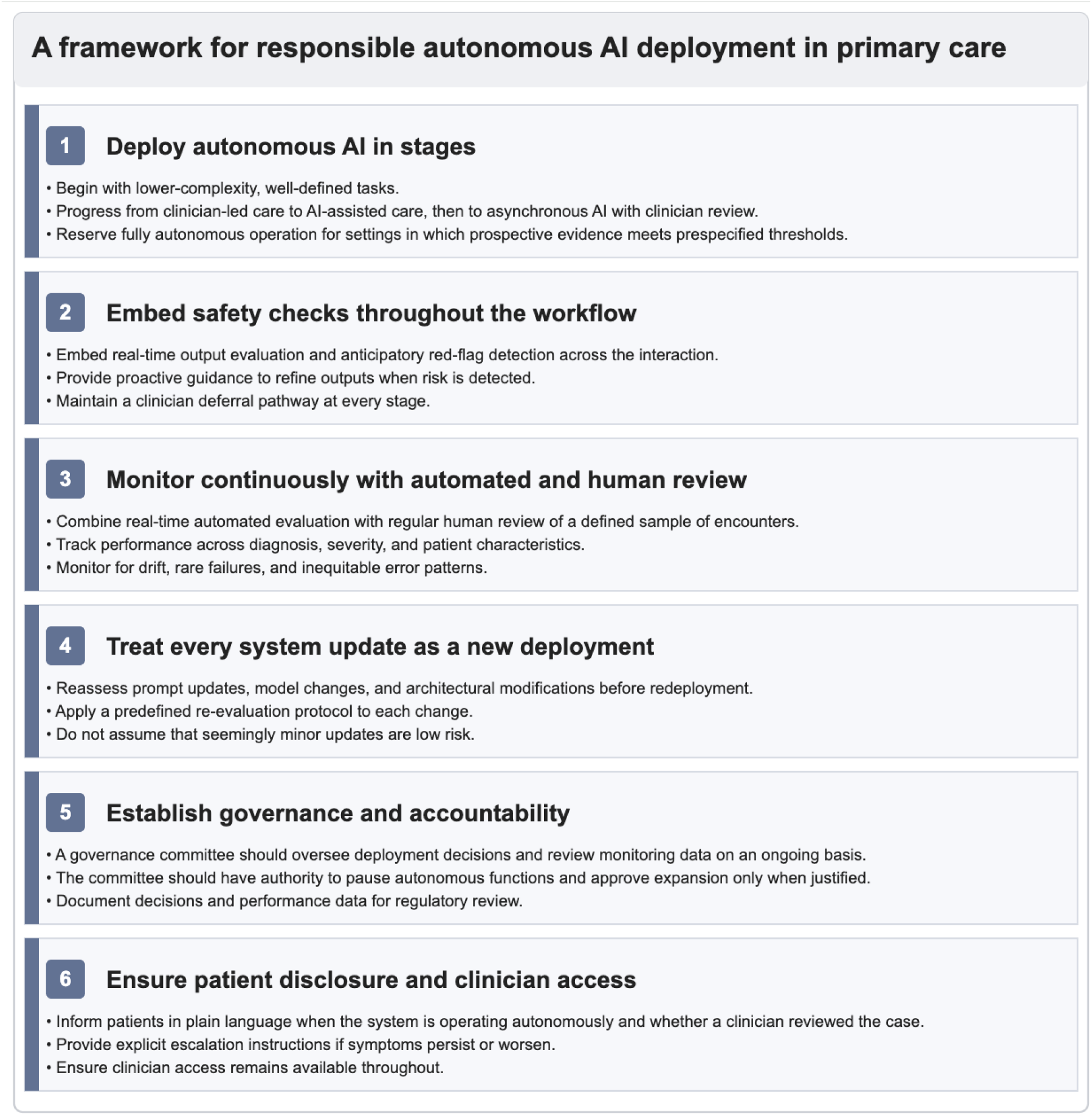
A framework for responsible autonomous AI deployment in Primary Care.

**Figure 5:**
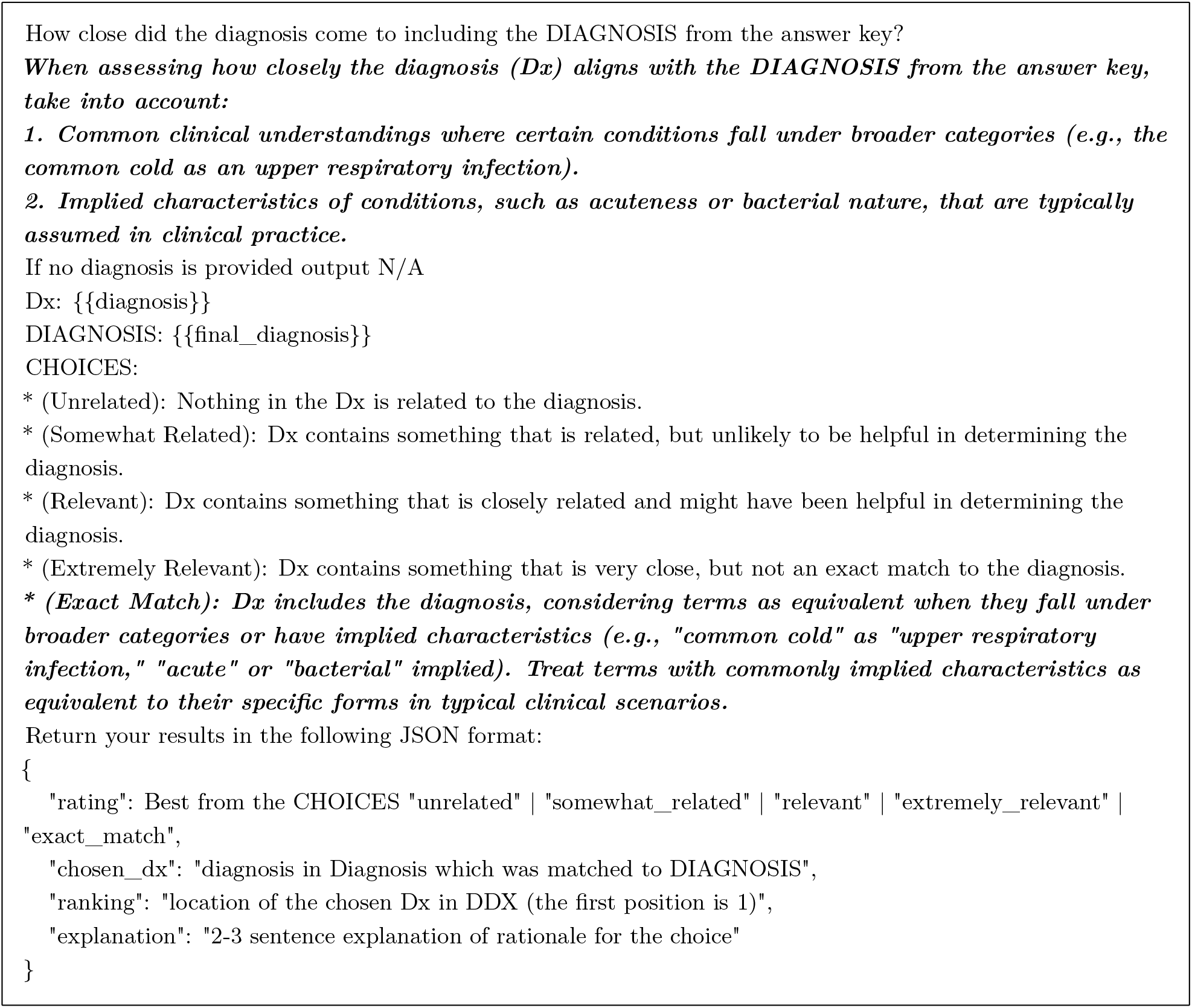
The prompt was adapted from Tu et al.^4^; modifications to account for broader diagnostic categories and implied clinical characteristics are shown in bold.

The path forward is not a binary transition from human oversight to machine autonomy, but a structured, evidence-driven expansion of the tasks for which autonomous operation has been validated. This evidence must be collected through studies like ours that use real-world clinical data and not curated groups of patients.

## 6. Conclusion

In a real-world primary care telemedicine setting, our multi-agent system, integrated in a safety architecture, demonstrated strong concordance with clinician-adjudicated disposition decisions and high diagnostic agreement within a clinician-led workflow, particularly for low-complexity cases. These findings support the evaluation and deployment of AI as comprehensive, end-to-end clinical systems featuring explicit safety gating, escalation, and deferral pathways, rather than as standalone models. To further define the appropriate scope of autonomous operation and track performance over time, larger prospective studies incorporating downstream clinical outcomes, subgroup analyses, and version-specific monitoring are warranted. We welcome collaboration with health institutions interested in advancing this work.

The question is no longer whether patients will seek autonomous AI for medical guidance, they already do at scale and outside established clinical frameworks. The real challenge is ensuring that the systems they rely on are held to the standards they deserve: evaluated on real-world encounters rather than simulated vignettes, assessed as integrated systems rather than isolated models, and governed by frameworks designed specifically for this technology. The future is not a binary shift from human oversight to machine autonomy, but a structured, evidence-based expansion of tasks for which autonomous operation has been prospectively validated. This approach enables honest risk assessment and paves the way for safer, more accessible, and more equitable healthcare.

## 7. Limitations of the study

Several limitations warrant consideration. First, the symptom checker disposition analysis was based on 161 adjudicated encounters, which limited the precision of performance estimates, particularly for less common error types and subgroup-specific analyses. Second, the diagnostic reference standard was the treating clinician’s documented diagnosis in the medical record rather than adjudication by a consensus panel. Although this approach reflects real-world clinical decision-making under routine care, including diagnoses entered by licensed clinicians responsible for patient management, it may also introduce variability related to documentation practices, diagnostic specificity, and individual clinical judgment. Third, diagnostic concordance was assessed with a structured evaluation prompt adapted from prior work rather than by universal physician adjudication. Although this approach enabled scalable review, it may not have fully captured all clinically meaningful distinctions or equivalences between diagnoses. Finally, although the platform serves a geographically diverse U.S. patient population, the findings should be evaluated further in international health care settings, where disease prevalence, care-seeking behavior, clinical workflows, and regulatory environments may differ.

## 8. Ethics Statement

This work originated as a quality-improvement initiative based on retrospective analysis of routinely collected clinical encounter data from a telemedicine platform. The study was reviewed by the Ethical AI Governance Committee of Curai Health, which deemed it exempt. The analyses reported in this manuscript used data generated during routine care, and no additional patient contact or study-specific intervention was undertaken. All data were anonymized (scrubbed of any patient revealing information) Users of the platform are informed through the platform’s privacy notice that their information may be used to provide services, improve the platform, and support research and quality-improvement activities.

## 9. Data availability

The data underlying this study are derived from real-world patient encounters and contain protected health information. Due to privacy and contractual restrictions, the underlying transcripts and associated encounter-level data cannot be shared publicly or on request. The structured evaluation prompt used for diagnostic concordance assessment is provided in the Supplementary Appendix.

## 10. Code availability

The system evaluated in this study is a proprietary clinical platform and the underlying code is not publicly available.

## Appendix

**Prompt used to evaluate diagnosis concordance**

